# COVID-19: Public Compliance with and Public Support for Stay-at-Home Mitigation Strategies

**DOI:** 10.1101/2020.04.22.20076141

**Authors:** Mark É. Czeisler, Mark E. Howard, Rebecca Robbins, Laura K. Barger, Elise R. Facer-Childs, Shantha M. W. Rajaratnam, Charles A. Czeisler

## Abstract

**Objectives:** Governments worldwide have recommended unprecedented measures to mitigate the coronavirus disease 2019 (COVID-19) pandemic. As pressure mounts to scale back these measures, understanding public compliance with and priorities for COVID-19 mitigation is critical. The main aim of this study was to assess public compliance with and support for government-imposed stay-at-home orders in nations and cities with different COVID-19 infection and death rates.

**Design:** In this cross-sectional study, questionnaires were administered to nationally representative respondents from April 2-8, 2020.

**Setting:** Regions with different disease prevalence included two nations [the United States (US—high) and Australia (AU—low)] and two cities [New York (NY—high) and Los Angeles (LA—low)].

**Participants:** For adults 18 years or older residing in specified regions, eligible respondents were empaneled until representative quotas were reached for age, gender, and either race and ethnicity (US, NY, LA) or ancestry (AU), matching the 2010 US or 2016 AU census. Of 8718 eligible potential respondents, 5573 (response rate, 63.9%) completed surveys (US: 3010; NY: 507; LA: 525; AU: 1531). The median age was 47 years (range, 18-89); 3039 (54.5%) were female.

**Exposure:** The prevalence of COVID-19 in each region (cumulative infections, deaths) as of April 8, 2020: US (458610, 15659), AU (5956, 45),^1^ NY (81803, 4571), LA (7530, 198).^2^

**Main Outcomes Measures:** Public compliance with and attitudes regarding government-imposed stay-at-home orders were evaluated and compared between regions.

**Results:** Of 5573 total respondents, 4560 (81.8%) reported compliance with recommended quarantine or stay-at-home policies (range of samples, 75.5%-88.2%). Despite significant disruptions of social and work life, health, and behavior, 5022 respondents (90.1%) supported government-imposed stay-at-home orders (range of samples, 88.9%-93.1%). Of these, 90.8% believe orders should last at least three more weeks or until public health or government officials recommend, with such support spanning the political spectrum.

**Conclusions:** Public compliance with stringent quarantine and stay-at-home policies was very high, in both highly-affected (US, NY) and minimally-affected regions (AU, LA). Despite extensive disruption of respondents’ lives, the vast majority supported continuation of long-term government-imposed stay-at-home orders. These findings have important implications for policymakers grappling with the decision as to when to lift restrictions.

## Introduction

To date, more than 2,500,000 confirmed cases and 175,000 deaths have been attributed to the novel coronavirus disease 2019 (COVID-19) pandemic.^3^ Absent widespread testing or safe and efficacious treatments, isolation and quarantine have been recommended worldwide for the first time in a century. Disease prevalence and associated public health policies have varied across jurisdictions and changed over time, largely without systematic assessment of public responses to the crisis or the mitigation strategies.

## Methods

### Study Design and Recruitment

To evaluate public compliance with and support for recommended COVID-19 mitigation strategies, we collected cross-sectional surveys of nationally representative respondents using demographic quota sampling.^4^ Surveys were administered to an online respondent panel by Qualtrics, LLC (Provo, Utah, and Seattle, Washington, US), a commercial survey company with a network of participant pools consisting of hundreds of suppliers. Recruitment methodologies include digital advertisements and promotions, word of mouth and membership referrals, social networks, TV and radio advertisements, and offline mail-based approaches.

Between April 2-8 2020 (a one-week period), samples were drawn from regions with markedly different infection and death rates from COVID-19 (Table 1), including nationwide samples in the United States (US, high incidence) and Australia (AU, low incidence), and two citywide samples in the New York (NY, high incidence) and Los Angeles (LA, low incidence) metropolitan areas.

**Table 1.**
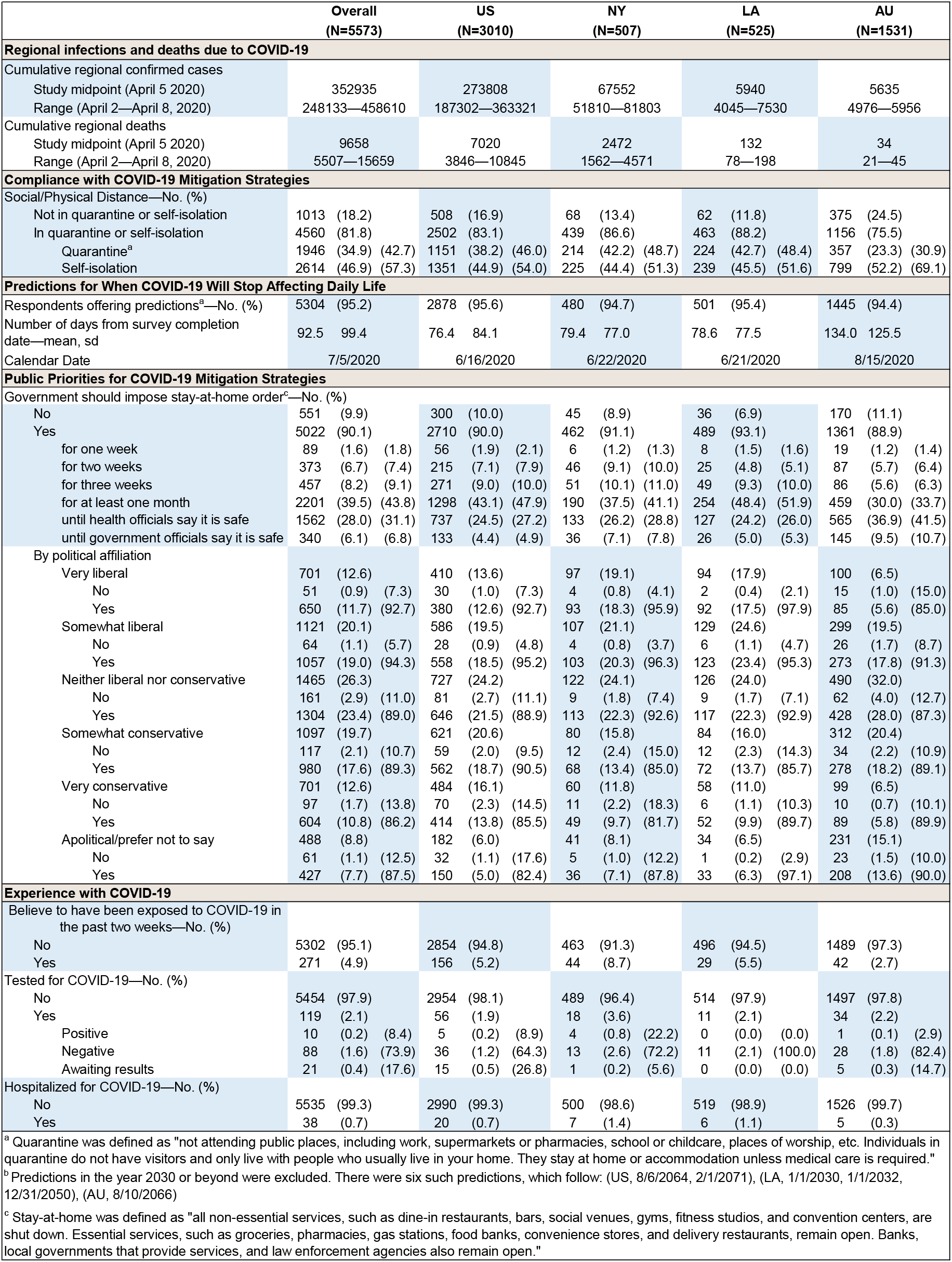
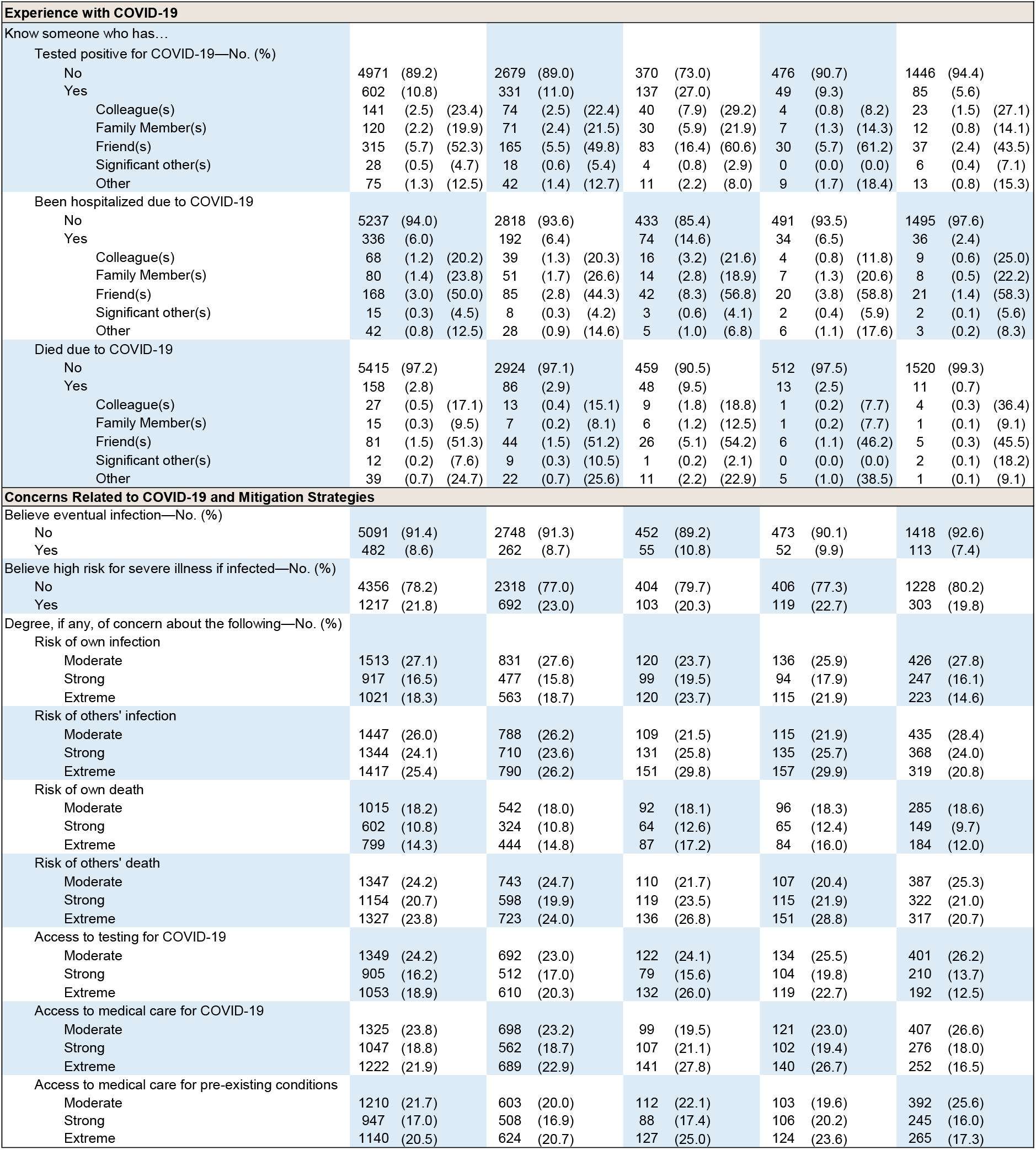
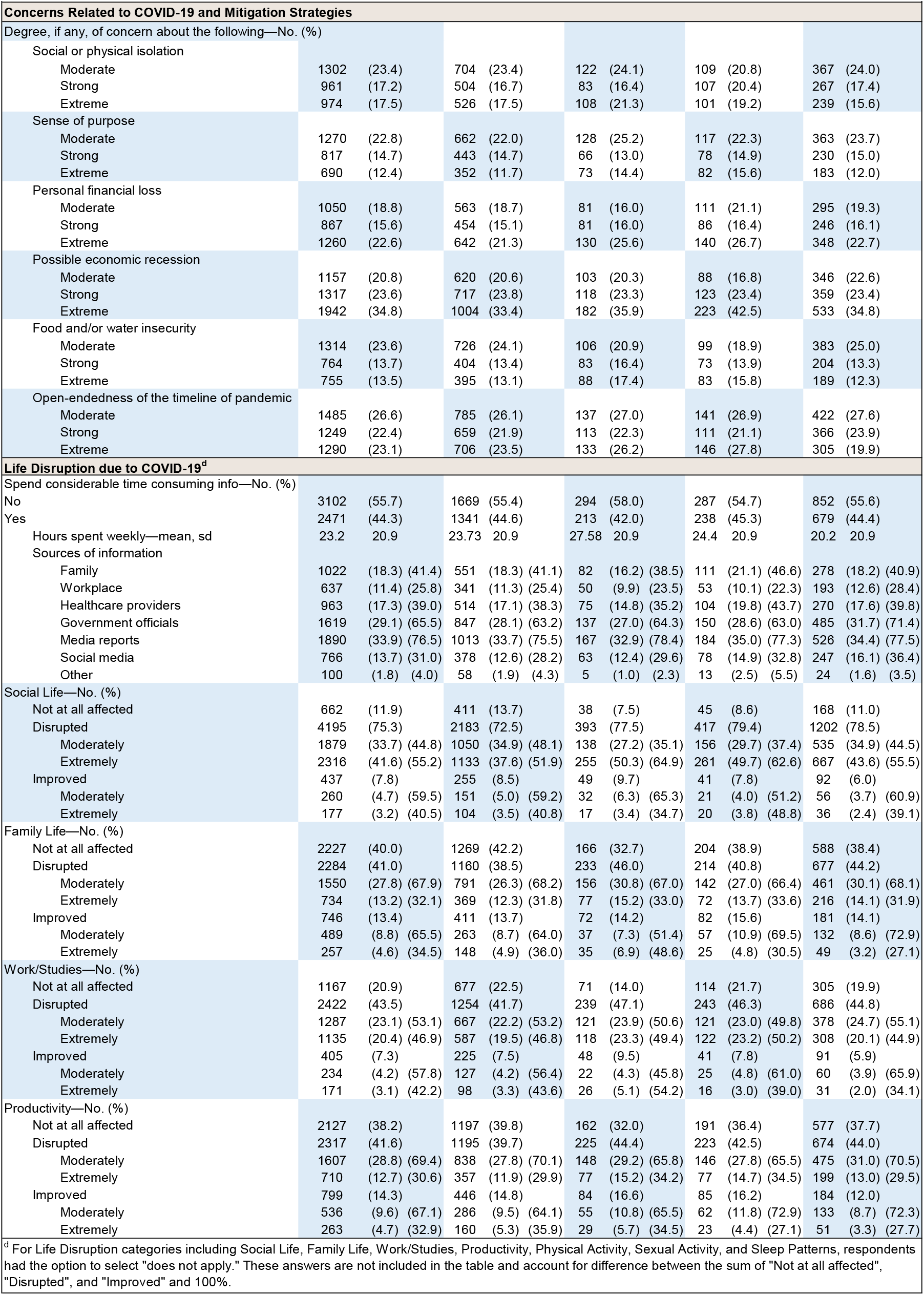
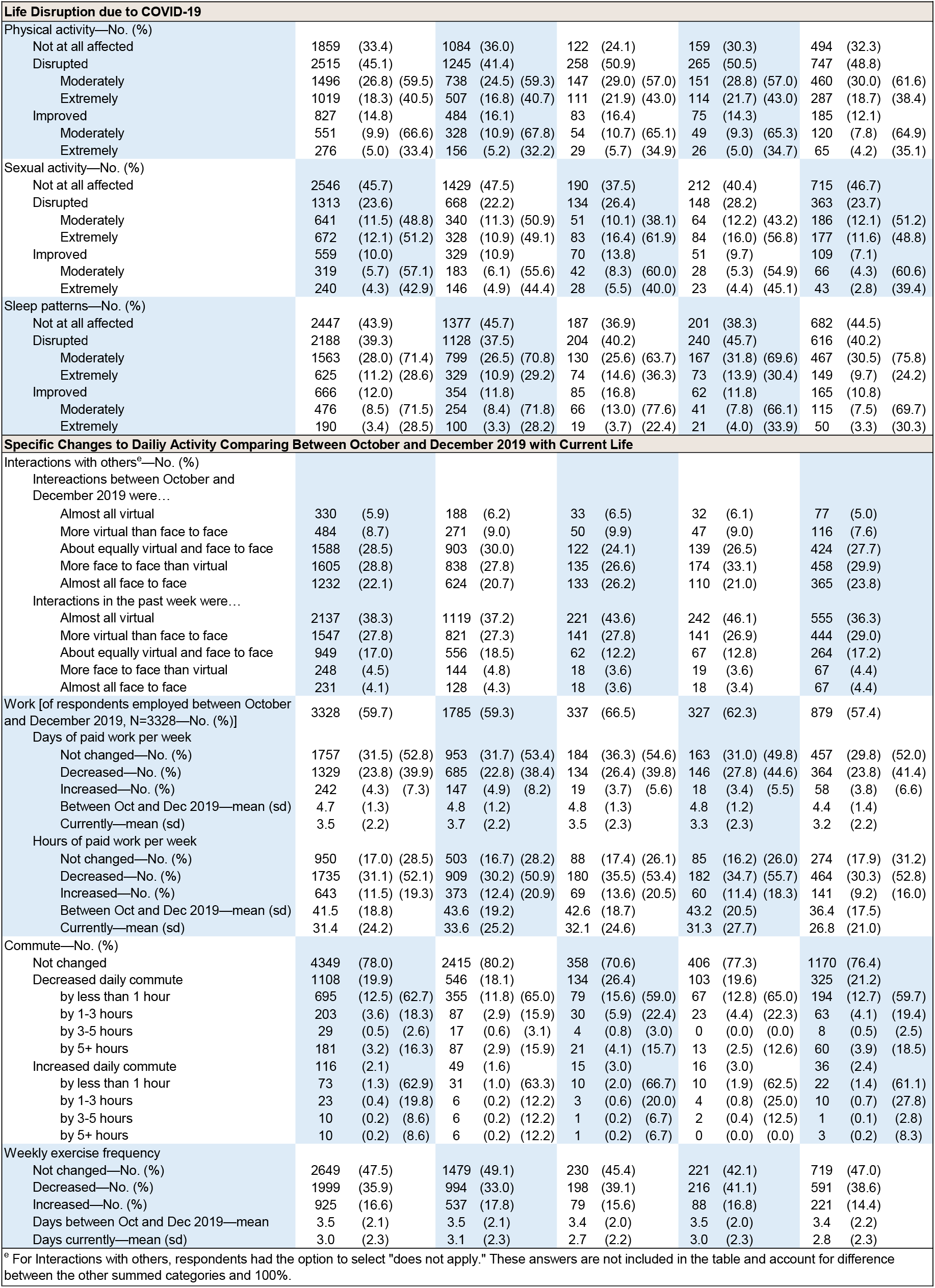
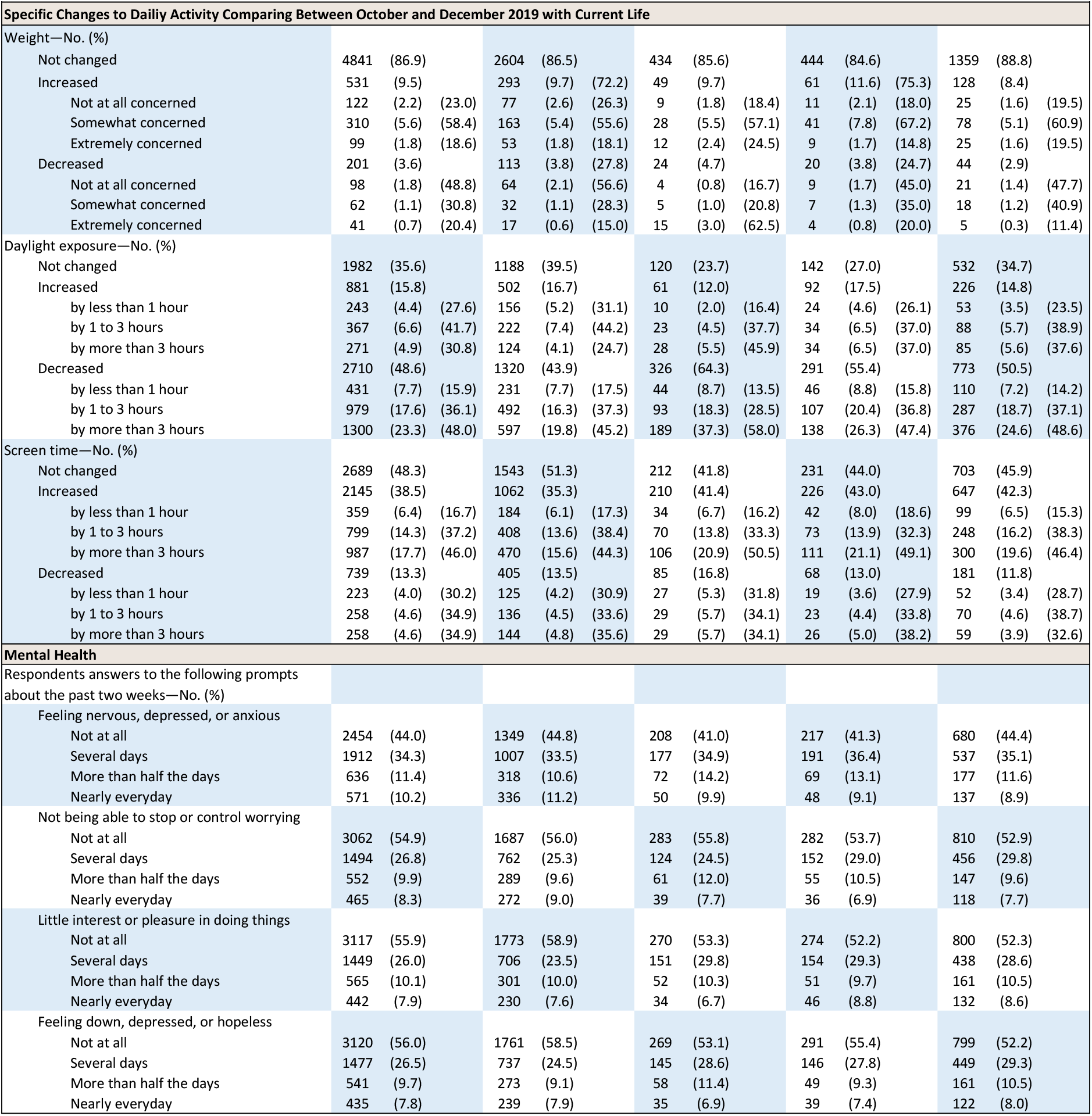
Summary of Results Overall and by Region. Survey responses are reported using descriptive statistics as indicated, including number of respondents (No.), percentage of respondents within a region (%), mean, and standard deviation (sd). For cases in which subgroups are stratified, the percentage of the total sample is reported first, and then percentage of the subgroup is reported in the second parentheses. For cases in which subgroups are not stratified, or where means and standard deviations are reported, the to the right is left blank. For the “Concerns Related to COVID-19 and Mitigation Strategies” section, only respondents rating moderate to extreme concern are reported (respondents reporting no or slight concern, or that the concern does not apply, are not shown). Country-level cumulative cases and deaths for US and AU were retrieved from World Health Organization COVID-19 Situational Reports.^1,28-29^ City-level cumulative cases and deaths for NY and LA were retrieved from The New York Times Coronavirus (Covid-19) Data in the United States project, based on reports from state and local health agencies.^2^

### Study Approval

The study protocol was approved by the Monash University Human Research Ethics Committee (MUHREC) and conducted in accordance with ethical guidelines. Respondents were informed of the study purposes and provided electronic consent prior to commencement. Investigators received anonymized responses.

### Population

Target numbers of respondent-completed surveys follows: US (3000), NY (500), LA (500), AU (1500). To be eligible to participate, respondents were required to report being aged 18 years or older with current residence in specified regions. Demographic sampling quotas were implemented for age, gender, and either race and ethnicity (US, NY, LA) or ancestry (AU), based on 2010 US and 2016 Australian census data. Potential respondents likely to qualify based on demographic characteristics listed in their Qualtrics panelist profile were targeted during recruitment; demographic questions were included in the survey to confirm eligibility. Potential respondents received invitations and could opt to participate by activating a survey link directing them to the participant information and consent page preceding the survey. Ineligible respondents who did not meet inclusion criteria (e.g., less than 18 years of age, not a resident of a targeted region) or exceeded set quotas (i.e., maximum demographic characteristic quota already met) were disempaneled from the survey.

### Survey Instruments

The surveys contained 86 [United States (US), New York (NY), Los Angeles (LA)] or 85 [culturally adapted for Australia (AU)] items, with each item requiring a response, and was designed to take approximately 15 minutes to complete. Respondents were required to self-report demographic characteristics and respond to questions about COVID-19 and mitigation strategies including compliance, priorities, sources of concern, and comparisons of current lifestyle versus lifestyle between October and December 2019 (i.e., before COVID-19 and mitigation strategies). Additional health-related questions were asked independent of COVID-19. When possible, brief validated instruments were used, including the Short-Form Sleep Condition Indicator (SCI-01) for insomnia risk assessment, Patient Health Questionnaire-4 (PHQ-4) for anxiety and depression risk assessment, the Perceived Stress Scale-4 (PSS-4) for perceived stress assessment, and the Mini Z for burnout risk assessment.^5-8^ When required, validated instruments were adapted, including the Horne and Östberg Morningness-Eveningness Questionnaire (MEQ) for chronotype assessment, the μshort Munich ChronoType Questionnaire (μMCTQ) for chronotype and sleep behavior assessment, Obstructive Sleep Apnoea 50 (OSA50) for obstructive sleep apnea risk assessment, single-item physical activity measure, and Hurt-Insult-Threaten-Scream (HITS) screening tool for domestic violence.^9-14^

### Quality Screening

To verify response quality, Qualtrics conducted standardized quality screening and data scrubbing procedures. Techniques included algorithmic analysis for attention patterns, click-through behavior, duplicate responses, keystroke analysis, machine responses, and inattentiveness. Country-specific geolocation verification via IP address mapping was used to ensure respondents were from the country specified in their response. Respondents who failed an attention or speed check, along with any responses identified by the data scrubbing algorithms, were excluded from the final sample.

## Results

Between April 2 and April 8, 2020, respondents completed 5573 surveys (US: 3010; NY: 507; LA: 525; AU: 1531) with a 63.9% response rate (Figure 1). Overall, 3039 (54.5%) were female; the median age of participants was 47 years (range, 18-89). See Table 2 for respondent characteristics. Cross-sectional results of COVID-19-mitigation compliance, public priorities, and life impact for each sample are reported by region (Figure 2; Table 1). Altogether, 4560 respondents (81.8%) reported quarantining or voluntarily self-isolating (range of samples, 75.5%-88.2%). Moreover, 5022 (90.1%) believe a government-imposed stay-at-home order is warranted (range of samples, 88.9%-93.1%). Of these, 90.8% believe the order should last at least three more weeks (9.1%), a month or longer (43.8%), or until public health (31.1%) or government officials (6.8%) determine it is safe to lift the restrictions. Of 5304 respondents (95.2%) who made predictions, the average predicted date by which COVID-19 would no longer affect their daily lives was between mid-June and mid-August, though there was high variability in predictions. Strong support for a government-imposed stay-at-home order spanned the political spectrum (Table 1).

**Table 2.** Self-Reported Respondent Characteristics. Self-reported characteristics overall and in quota samples collected in the US, NY, LA, and AU. For age, mean (standard deviation), median, and range are shown per sample. For all other characteristics, the number and percentage of respondents are reported by cohort. Race and ethnicity (US, NY, LA) or ancestry (AU) were reported in based on questions culturally adapted to match the characteristic data collected in the 2010 United States and 2016 Australian Census, respectively.

| Characteristic | Overall<br>(N=5573) | US<br>(N=3010) | NY<br>(N=507) | LA<br>(N=525) | (N=1531) |
| --- | --- | --- | --- | --- | --- |
| Age (years) |  |  |  |  |  |
| Mean (SD) | 47.1 (17.3) | 47.4 (16.9) | 46.7 (18.0) | 45.5 (17.0) | 45.6 (17.3) |
| Median | 47 | 48 | 45 | 45 | 44.5 |
| Range | 18-89 | 18-89 | 18-86 | 18-87 | 18-89 |
| Gender—No. (%) |  |  |  |  |  |
| Female | 3039 (54.5) | 1683 (55.9) | 239 (47.1) | 275 (52.4) | 842 (55.0) |
| Male | 2530 (45.4) | 1325 (44.0) | 268 (52.9) | 250 (47.6) | 687 (44.9) |
| Other | 4 (0.1) | 2 (0.1) | 0 (0.0) | 0 (0.0) | 2 (0.1) |
| Race<sup>a</sup> (All US, N=4042)—No. (%) |  |  |  |  |  |
| White | 3196 (79.1) | 2423 (80.5) | 373 (73.6) | 400 (76.2) |  |
| Black or African American | 428 (10.6) | 313 (10.4) | 63 (12.4) | 52 (9.9) |  |
| Asian | 256 (6.3) | 192 (6.4) | 32 (6.3) | 32 (6.1) |  |
| American Indian or Alaskan Native | 80 (2.0) | 60 (2.0) | 9 (1.8) | 11 (2.1) |  |
| Native Hawaiian or Pacific Islander | 22 (0.5) | 17 (0.6) | 3 (0.6) | 2 (0.4) |  |
| Other | 182 (4.5) | 104 (3.5) | 38 (7.5) | 40 (7.6) |  |
| Ethnicity (All US, N=4042)—No. (%) |  |  |  |  |  |
| Hispanic or Latino | 424 (10.5) | 265 (8.8) | 69 (13.6) | 90 (17.1) |  |
| Not Hispanic or Latino | 3618 (89.5) | 2745 (91.2) | 438 (86.4) | 435 (82.9) |  |
| Ancestry<sup>b</sup> (AU, N=1531)—No. (%) |  |  |  |  |  |
| Australian |  |  |  |  | 556 (36.3) |
| English |  |  |  |  | 501 (32.7) |
| Other European (Irish, Scottish, German, Italian, Greek, Dutch) |  |  |  |  | 346 (22.6) |
| Scottish |  |  |  |  | 95 (6.2) |
| Chinese |  |  |  |  | 90 (5.9) |
| Indian |  |  |  |  | 45 (2.9) |
| Indigenous—Aboriginal Australians and Torres Strait Islanders |  |  |  |  | 16 (1.0) |
| Other |  |  |  |  | 455 (29.8) |
| Highest degree or level of education completed—No. (%) |  |  |  |  |  |
| Less than high school | 107 (1.9) | 61 (2.0) | 4 (0.8) | 5 (1.0) | 37 (2.4) |
| High school or equivalent | 1257 (22.6) | 524 (17.4) | 81 (16.0) | 61 (11.6) | 591 (38.6) |
| Some college | 1444 (25.9) | 910 (30.2) | 121 (23.9) | 157 (29.9) | 256 (16.7) |
| Bachelor's degree (4-year) or equivalent | 1806 (32.4) | 927 (30.8) | 159 (31.4) | 212 (40.4) | 508 (33.2) |
| Doctoral or professional degree | 917 (16.5) | 567 (18.8) | 137 (27.0) | 88 (16.8) | 125 (8.2) |
| Prefer not to say | 42 (0.8) | 21 (0.7) | 5 (1.0) | 2 (0.4) | 14 (0.9) |
| <sup>a</sup> For the US sample, respondents had the option to select more than one racial affiliation. |  |  |  |  |  |
| <sup>b</sup> For the AU sample, respondents had the option to select up to two ancestral affiliations. The 'Other' category includes Filipino, Vietnamese, Lebanese, Hmong, Kurdish, Maori, and Australian South Sea Islander. |  |  |  |  |  |

**Table 2.** Self-reported characteristics overall and in quota samples collected in the US, NY, LA, and AU.
| Characteristic | Overall<br>(N=5573) | US<br>(N=3010) | NY<br>(N=507) | LA<br>(N=525) | AU<br>(N=1531) |
| --- | --- | --- | --- | --- | --- |
| Marital status—No. (%) |  |  |  |  |  |
| Married | 2724 (48.9) | 1567 (52.1) | 231 (45.6) | 226 (43.0) | 700 (45.7) |
| Living with partner | 533 (9.6) | 241 (8.0) | 43 (8.5) | 51 (9.7) | 198 (12.9) |
| Separated | 92 (1.7) | 32 (1.1) | 7 (1.4) | 2 (0.4) | 51 (3.3) |
| Divorced | 490 (8.8) | 291 (9.7) | 40 (7.9) | 46 (8.8) | 113 (7.4) |
| Widowed | 178 (3.2) | 109 (3.6) | 12 (2.4) | 21 (4.0) | 36 (2.4) |
| Never married | 1490 (26.7) | 739 (24.6) | 165 (32.5) | 169 (32.2) | 417 (27.2) |
| Prefer not to say | 66 (1.2) | 31 (1.0) | 9 (1.8) | 10 (1.9) | 16 (1.0) |
| 2019 household income (USD)—No. (%) |  |  |  |  |  |
| Less than \$24,999 | 940 (16.9) | 454 (15.1) | 57 (11.2) | 67 (12.8) | 362 (23.6) |
| \$25,000 to \$49,999 | 1296 (23.3) | 641 (21.3) | 88 (17.4) | 88 (16.8) | 479 (31.3) |
| \$50,000 to \$99,999 | 1723 (30.9) | 989 (32.9) | 139 (27.4) | 164 (31.2) | 431 (28.2) |
| \$100,000 to \$199,999 | 1054 (18.9) | 657 (21.8) | 151 (29.8) | 134 (25.5) | 112 (7.3) |
| More than \$200,000 | 229 (4.1) | 132 (4.4) | 41 (8.1) | 42 (8.0) | 14 (0.9) |
| Prefer not to say | 331 (5.9) | 137 (4.6) | 31 (6.1) | 30 (5.7) | 133 (8.7) |
| 2019 employment status—No. (%) |  |  |  |  |  |
| Employed full-time | 2245 (40.3) | 1284 (42.7) | 246 (48.5) | 217 (41.3) | 498 (32.5) |
| Employed part-time | 760 (13.6) | 338 (11.2) | 63 (12.4) | 61 (11.6) | 298 (19.5) |
| Self-employed | 361 (6.5) | 189 (6.3) | 30 (5.9) | 52 (9.9) | 90 (5.9) |
| Student | 337 (6.0) | 147 (4.9) | 30 (5.9) | 36 (6.9) | 124 (8.1) |
| Retired | 1268 (22.8) | 734 (24.4) | 101 (19.9) | 110 (21.0) | 323 (21.1) |
| Unemployed | 714 (12.8) | 384 (12.8) | 45 (8.9) | 55 (10.5) | 230 (15.0) |
| Political ideology—No. (%) |  |  |  |  |  |
| Very liberal | 701 (12.6) | 410 (13.6) | 97 (19.1) | 94 (17.9) | 100 (6.5) |
| Slightly liberal | 1121 (20.1) | 586 (19.5) | 107 (21.1) | 129 (24.6) | 299 (19.5) |
| Neither liberal nor conservative | 1465 (26.3) | 727 (24.2) | 122 (24.1) | 126 (24.0) | 490 (32.0) |
| Slightly conservative | 1097 (19.7) | 621 (20.6) | 80 (15.8) | 84 (16.0) | 312 (20.4) |
| Very conservative | 701 (12.6) | 484 (16.1) | 60 (11.8) | 58 (11.0) | 99 (6.5) |
| Apolitical and/or prefer not to say | 488 (8.8) | 182 (6.0) | 41 (8.1) | 34 (6.5) | 231 (15.1) |

**Figure 1.**
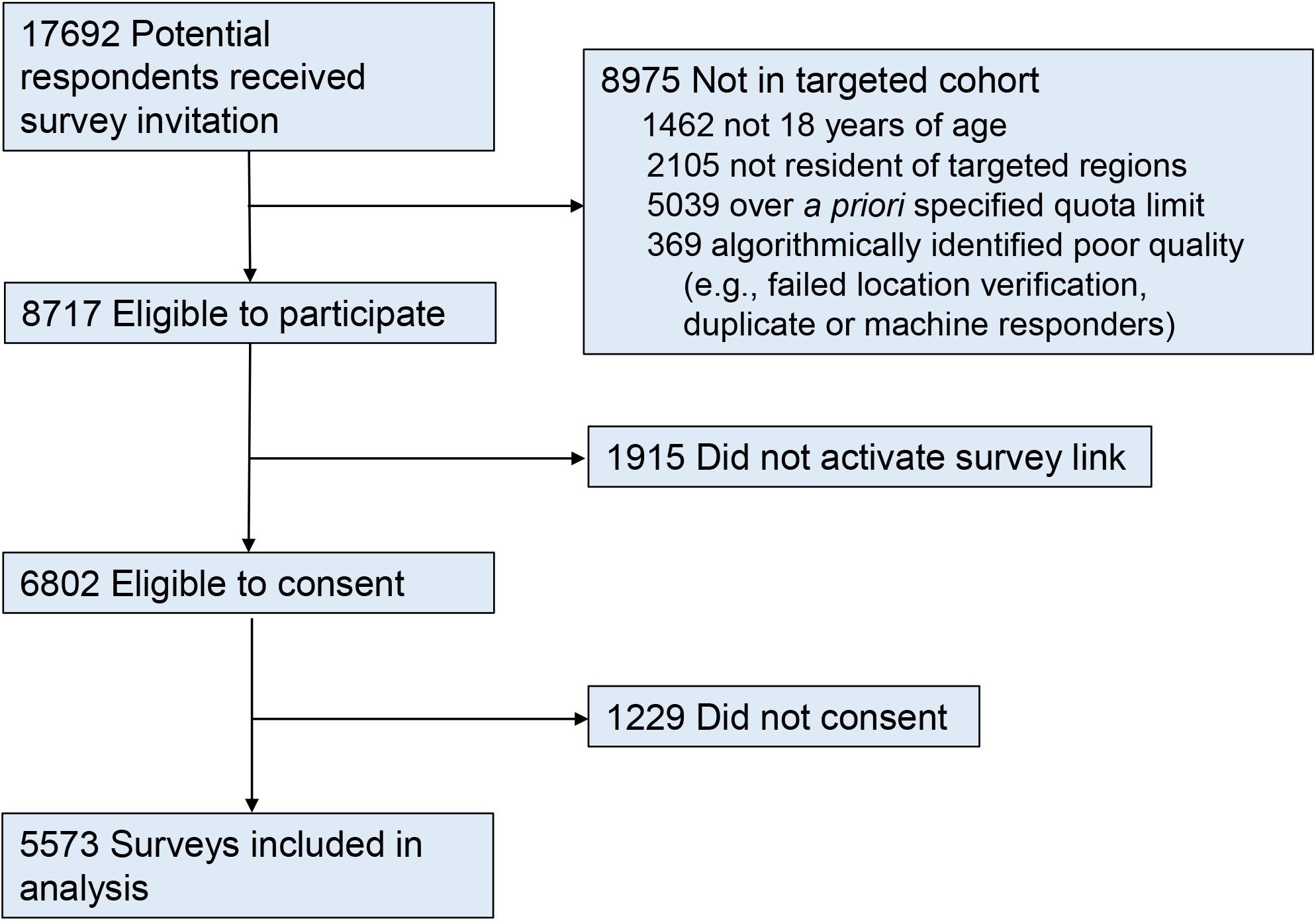
Flow of Survey Respondents. The survey was managed through an online respondent panel by Qualtrics. *A priori* quota limits were determined prior to study initiation to ensure nationally representative samples were collected and included the following: age, gender, and either race and ethnicity (US, NY, LA) or ancestry (AU), based on 2010 US and 2016 Australian census data. Of 8718 eligible potential respondents, 5573 completed surveys, providing a 63.9% response rate.

**Figure 2.**
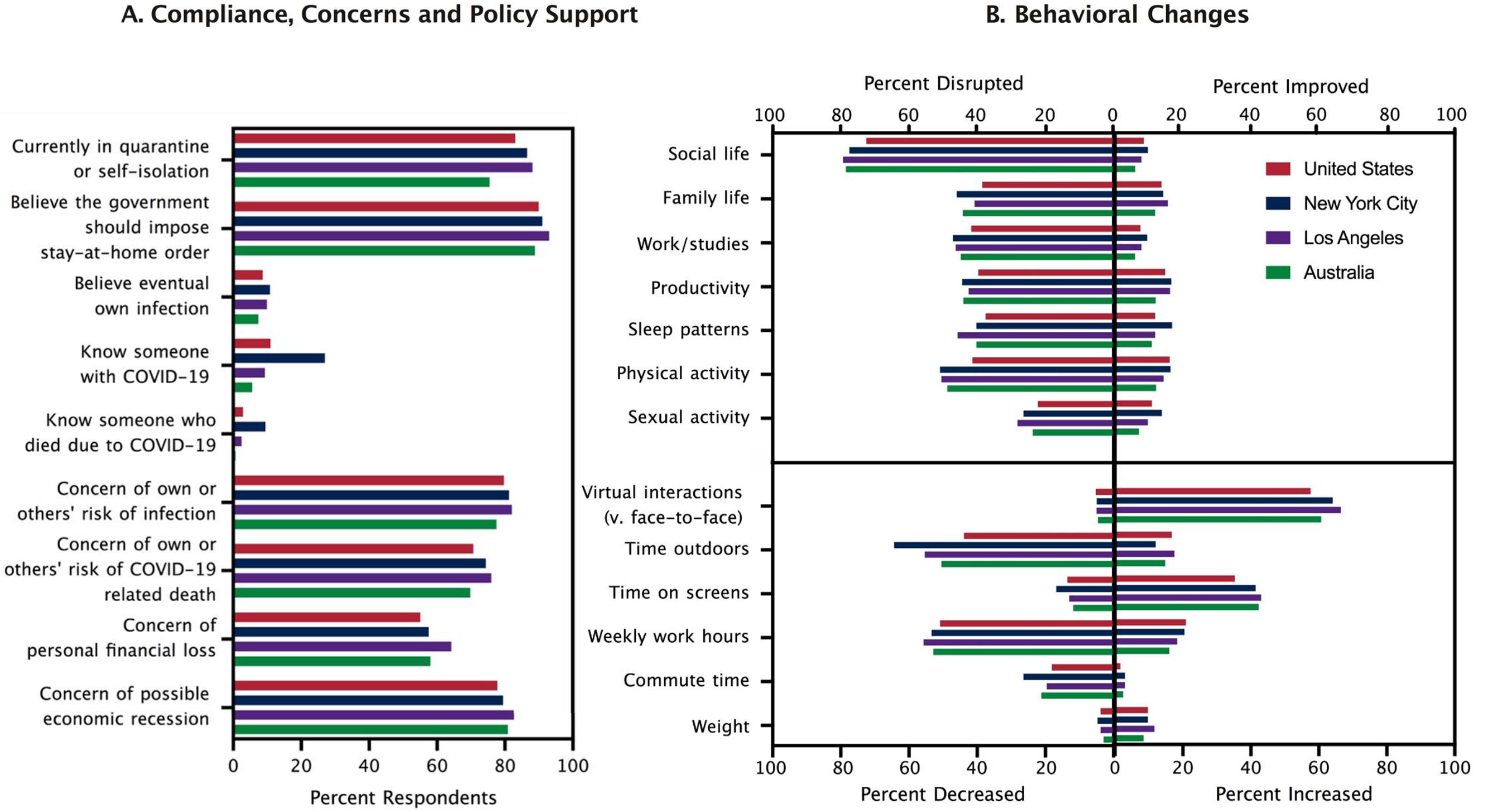
Public COVID-19 Mitigation Compliance, Concerns, Policy Support and Experience (A) Percentage of respondents by region who report: compliance with nationally recommended mitigation strategies; support for a government-mandated stay-at-home order; perceived risk of eventual infection with COVID-19; personal experience with COVID-19 (i.e., knowing someone who was infected with or died from COVID-19); and moderate to extreme concerns regarding: one’s own or others’ risk of infection with or death from COVID-19, personal financial loss, and possible economic recession. (B) Upper quadrants: Impact by region of COVID-19 and mitigation strategies on social life, family life, work and/or study, productivity, sleep patterns, physical activity, and sexual activity; percentage of respondents reporting that the indicated behavioral category was moderately to extremely disrupted or improved is shown. Lower quadrants: Percentage of respondents reporting decreases or increases in six categories [virtual interactions vs. face-to-face interactions; time spent outdoors during daylight hours; time on light-emitting screens; weekly work hours (among respondents employed in the fourth quarter of 2019, n=3328); commute time; and weight] at the time of the survey in April, 2020 (after the COVID-19 pandemic was declared and mitigation was underway) as compared to the fourth quarter of 2019 (before the COVID-19 pandemic was declared).

Overall, 4431 respondents (79.5%, range of samples, 77.5%-82.1%) reported moderate to extreme concern about their own (61.9%) or others’ (75.5%) infection and 3974 (71.3%, range of samples, 69.8%-76.0%) about their own (43.4%) or others’ (68.7%) death. Access to testing (59.3%), medical care for COVID-19 (64.5%), medical care for pre-existing conditions due to hospital overload (59.2%), social or physical isolation (58.1%), and sense of purpose (49.8%) were also sources of moderate to extreme concern. Overall, 1217 respondents (21.8%) identified as high risk for severe COVID-19 infection. Across regions, nearly half (42.0%-45.3%) reported spending considerable time (average, 23.2 hours per week) consuming information (media, government reports, health officials, family) about COVID-19. NY had the highest percentage of respondents who reported knowing someone who has tested positive for (27.0% vs. 5.6%-11.0%), been hospitalized for (14.6% vs. 2.4%-6.5%) or died from (9.5% vs. 0.7%-2.9%) COVID-19.

Consistent across regions, respondents reported that COVID-19 and mitigation strategies have caused moderate to extreme disruption of social life (75.3%), family life (41.0%), work/studies (43.5%), productivity (41.6%), physical activity (45.1%), sexual activity (23.6%), and sleep patterns (39.3%). Overall, 1999 respondents (35.9%) reported exercising less frequently, and 409 (7.4%) reported concerning weight gain. Daily outdoor light exposure was reduced by 1 hour or more in 2279 respondents (40.9%). The estimated percentage of virtual interactions (versus face-to-face) increased from 14.6% to 66.1%, and 1786 respondents (32.0%) reported more than 1 hour increase in daily screen time.

Widespread concerns included the possibility of an economic recession and open-endedness of COVID-19 and mitigation measures (79.2% and 72.2%, respectively). A total of 3119 respondents (56.0%) reported feeling anxious or nervous, 2453 (44.0%) depressed or hopeless, and 2511 (45.1%) unable to stop or control worrying at least several days in the prior two weeks.

## Discussion

Resounding compliance with and support for disruptive mitigation measures evidenced in these nationally representative samples, despite belief by 91.4% of respondents that they will never be infected with COVID-19 (range of samples, 89.2%-92.6%), suggests that controlling COVID-19 is a top public priority.^15^ We used quota sample surveys to rapidly assess public compliance, priorities, and life impact related to COVID-19 and mitigation strategies. We recognize the potential for self-selection bias; however, the high response rate (63.9%) and consistency of responses across cities and countries despite different rates of infection, governments, and mitigation strategies indicate that these results are robust. Contrary to public attitudes and compliance with recommended mitigation during the last pandemic^16-17^ declared by the World Health Organization for novel influenza A (H1N1) in 2009,^18^ the public response to the COVID-19 pandemic represents a hitherto unprecedented and rapid level of compliance with public health emergency measures that have and will continue to have a profound impact on economics and public life. These results demonstrate an escalated public response compared to before shelter-in-place orders were widely implemented,^19^ and contribute to a growing body of evidence that mitigation strategies for COVID-19, like those for previous outbreaks, are associated with significant disruption of life and general health consequences.^20-25^ These findings may also provide insight into behavioral countermeasures related to sleep, exercise, and diet that may reduce adverse health consequences of COVID-19 mitigation measures. As controversies over the legality^26^ and balance between duration and nature of mitigation strategies and related consequences continue to mount, and with the recent prospect of repeated and protracted stay-at-home orders being recommended over the next two years,^27^ assessment of public priorities, compliance, and life impact is paramount.

Compliance with and support for COVID-19 mitigation strategies, alongside concerns and life impact, were assessed in nationally representative samples in the United States and Australia. These timely findings indicate that the public is not only willing to accept current mitigation measures and their associated costs, but that people endorse their continuation until the COVID-19 pandemic is controlled.

## Data Availability

The authors confirm that the data supporting the findings of this study are available within the article.

## Author Contributions

M.É.C. had full access to all data in the study and takes responsibility for the integrity of the data and accuracy of data analyses.

Concept and design: All authors.

Acquisition and analysis of data: M.É.C.

Drafting of the manuscript: M.É.C., S.M.W.R., C.A.C.

Critical revision of the manuscript for important intellectual content: All authors.

Supervision: M.E.H., S.M.W.R., C.A.C.

## Funders

This study was supported in part by the Institute for Breathing and Sleep, Austin Health; the Turner Institute for Brain and Mental Health, Monash University; and by a gift to the Harvard Medical School from Philips Respironics. M.É.C. was supported by a 2020 Fulbright Future Scholarship funded by the Kinghorn Foundation through the Australian-American Fulbright Commission. L.K.B. and C.A.C. were supported in part by the National Institute of Occupational Safety and Health R01-OH-010300. C.A.C. serves as the incumbent of an endowed professorship provided to Harvard Medical School by Cephalon, Inc. The funders were not involved in the design and conduct of the study, the collection, preparation, or interpretation of the data, or the preparation or approval of the manuscript.

## Conflict of Interest Disclosures

C.A.C. reports grants from Cephalon Inc., Jazz Pharmaceuticals Plc., Inc., Philips Respironics, Inc., Regeneron Pharmaceuticals, ResMed Foundation, San Francisco Bar Pilots, Sanofi S.A., Sanofi-Aventis, Inc, Schneider Inc., Sepracor, Inc, Mary Ann & Stanley Snider via Combined Jewish Philanthropies, Teva Pharmaceuticals Industries, Ltd.; and personal fees from Bose Corporation, Columbia River Bar Pilots, Ganésco Inc., Institute of Digital Media and Child Development, Klarman Family Foundation, Samsung Electronics, Quest Diagnostics, Inc., Teva Pharma Australia, Vanda Pharmaceuticals, Washington State Board of Pilotage Commissioners, Zurich Insurance Company, Ltd. In addition, C.A.C. holds a number of process patents in the field of sleep/circadian rhythms (e.g., photic resetting of the human circadian pacemaker) and holds an equity interest in Vanda Pharmaceuticals, Inc. Since 1985, C.A.C. has also served as an expert on various legal and technical cases related to sleep and/or circadian rhythms, including those involving the following commercial entities: Casper Sleep Inc., Comair/Delta Airlines, Complete General Construction Company, FedEx, Greyhound, HG Energy LLC, Purdue Pharma, LP, Steel Warehouse Inc., Stric-Lan Companies LLC, Texas Premier Resource LLC, and United Parcel Service (UPS). CAC receives royalties from Philips Respironics, Inc. for the Actiwatch-2 and Actiwatch-Spectrum devices. Interests for C.A.C. were reviewed and managed by Brigham and Women’s Hospital and Partners HealthCare in accordance with their conflict of interest policies.

## Additional Contributions

We thank Qualtrics, LLC for their support of survey administration and data collection.

## Notes

### Clinical Trial

Not applicable.

